# Establishment of an evaluation panel for the decentralized technical evaluation of the sensitivity of 31 rapid detection tests for SARS-CoV-2 diagnostics

**DOI:** 10.1101/2021.05.11.21257021

**Authors:** Andreas Puyskens, Eva Krause, Janine Michel, Micha Nübling, Heinrich Scheiblauer, Daniel Bourquain, Marica Grossegesse, Roman Valusenko, Viktor Corman, Christian Drosten, Katrin Zwirglmaier, Roman Wölfel, Constanze Lange, Jan Kramer, Johannes Friesen, Ralf Ignatius, Michael Müller, Jonas Schmidt-Chanasit, Petra Emmerich, Lars Schaade, Andreas Nitsche

## Abstract

**Background:** The detection of SARS-CoV-2 with rapid diagnostic tests has become an important tool to identify infected people and break infection chains. These rapid diagnostic tests are usually based on antigen detection in a lateral flow approach.

**Aims & Methods:** While for PCR diagnostics the validation of a PCR assay is well established, for antigen tests e.g. rapid diagnostic tests there is no common validation strategy. Here we present the establishment of a panel of 50 pooled clinical specimens that cover a SARS-CoV-2 concentration range from approximately 1.1 × 10^9^ to 420 genome copies per mL of specimen. The panel was used to evaluate 31 rapid diagnostic tests in up to 6 laboratories.

**Results:** Our results show that there is significant variation in the detection limits and the clinical sensitivity of different rapid diagnostic tests. We conclude that the best rapid diagnostic tests can be applied to reliably identify infectious individuals who are presenting with SARS-CoV-2 loads correlated to 10^6^ genome copies per mL of specimen. Infected individuals displaying SARS-CoV-2 genome loads corresponding to less than 10^6^ genome copies per mL will be identified by only some rapid diagnostics tests, while many tests miss these viral loads to a large extent.

**Conclusions:** Sensitive RDTs can be applied to identify infectious individuals with high viral loads, but not to identify infected individuals.

## Background

PCR-based diagnostics of SARS-CoV-2 is a well-established method with numerous commercially available kits and in-house assays published over the last months [1]. Although it is beyond question that in particular real-time PCR provides an unrivalled degree of analytical sensitivity and reproducibility, there are some obvious limitations [2]. First of all, PCR-based diagnostics requires a functioning laboratory infrastructure and skilled personnel. The PCR reaction itself requires only about 2 hours, including pre- and post-analytical steps; however, time to result in high-throughput mode is about 24 hours or more.

The need for faster and simpler approaches to diagnose a SARS-CoV-2 infection is therefore evident as well as the need for on-site tests and for diagnostics in regions of lower standards of laboratory infrastructure [3–5]. One promising technology is using lateral flow immunoassays detecting SARS-CoV-2-specific proteins in respiratory secretions which operate within less than 30 minutes (Rapid Diagnostic Test: RDT) [6]. The trade-off for a simple and quick diagnostic test is an often significantly lower analytical sensitivity and specificity compared to nucleic acid amplification techniques like PCR [7,8].

Beside the traditional RDTs whose read-out is performed visually, there are assays that utilize readers for the identification of positive signals. These readers can provide a better sensitivity, reproducibility and objectivity, in particular with fluorescence-based-formats; however, the mobility and parallel testing of many specimens can be negatively affected.

Here we describe the decentralized evaluation of the sensitivity of 31 RDTs by up to six German laboratories investigating an identical panel of 50 pools of clinical specimens. By this approach, we generated at least two independent results per RDT and hence addressed inter-laboratory variations that have to be considered when RDTs are performed in different locations by different persons.

## Material & Methods

### Evaluation panel

To enable the systematic and comparable decentralized evaluation of numerous RDTs, a panel of 50 samples was compiled by pooling in total approximately 500 specimens which had been collected between March and September 2020 (Panel 1V1). For this purpose, up to ten respiratory specimens obtained for routine diagnostics with different virus loads were pooled. Pools were frozen at −80 °C. Real-time PCR [(9)] was applied to determine the RNA load per pool. In vitro RNA (provided by WHO) as well as the quantitative reference material provided by INSTAND were used for quantification (https://www.instand-ev.de). Finally, the panel covered a range of SARS-CoV-2 RNA from 1.1×10^9^ genomes per mL down to 420 genomes per mL. The study obtained ethical approval by the Berliner Ärztekammer (Berlin Chamber of Physicians, Eth 20/40). When Panel 1V1 was used up, new pools were generated by diluting the same samples as for Panel 1V1 (except for 4 pools 1–4 that had to be constituted from new clinical specimens collected between October 2020 and January 2021), resulting in comparable virus loads as determined by PCR, and the panel was labelled Panel 1V2. Panel 1V1 was compared to Panel 1V2 with RDT #3 and RDT #31 and showed identical results. Panel 1V1 and Panel 1V2 have been extensively used to evaluate the sensitivity of 122 RDTs as described in the tandem publication by Scheiblauer et al.)

Previous studies revealed that a minimal RNA genome copy number of 10^6^ genome copies per mL of specimen represents an amount of infectious virus particles which is required for successful virus propagation in cell culture [9–12]. To correlate the pools to potential infectivity by a specimen, we subjected the pools with ≥10^6^ genome copies per mL, corresponding to a CT value <25, to cell culture. Confirmation of replication-competent SARS-CoV-2 was achieved by inoculation of VeroE6 cells with the respective pools. Pools containing infectious SARS-CoV-2 were subsequently titrated on VeroE6 cells. However, even if pools containing higher amounts of SARS-CoV-2 RNA showed generally higher titres than those with lower genome numbers, we observed no significant correlation between the genome load and the titre (data not shown).

The specifications of the 50 pools are listed in table 1; pools allowing SARS-CoV-2 propagation are marked in bold.

**Table 1:** Characteristics of the 50 pools constituting Panel 1V1 and Panel 1V2.

| Panel 1V1 |  |  | Panel 1V2 |  |  |
| --- | --- | --- | --- | --- | --- |
| Pool-Nr. | CT/5 μL of RNA | RNA subjected to test | Pool-Nr. | CT/5 μL of RNA | RNA subjected to test |
| 1 | **17.55** | 1.1E+07 | 1 | **17.31** | 1.31E+07 |
| 2 | **20.54** | 1.4E+06 | 2 | **19.08** | 3.87E+06 |
| 3 | 20.38 | 1.6E+06 | 3 | **19.62** | 2.67E+06 |
| 4 | **20.98** | 1.0E+06 | 4 | **20.61** | 1.35E+06 |
| 5 | **20.28** | 1.7E+06 | 5 | **20.60** | 1.36E+06 |
| 6 | **20.20** | 1.8E+06 | 6 | 21.21 | 8.96E+05 |
| 7 | **21.71** | 6.4E+05 | 7 | 22.15 | 4.70E+05 |
| 8 | 21.95 | 5.4E+05 | 8 | 22.32 | 4.18E+05 |
| 9 | 22.14 | 4.7E+05 | 9 | 23.13 | 2.39E+05 |
| 10 | 22.88 | 2.8E+05 | 10 | 23.21 | 2.27E+05 |
| 11 | 22.34 | 4.1E+05 | 11 | 23.13 | 2.27E+05 |
| 12 | **21.82** | 5.9E+05 | 12 | 22.12 | 4.79E+05 |
| 13 | 23.32 | 2.1E+05 | 13 | 25.29 | 5.42E+04 |
| 14 | 24.28 | 1.1E+05 | 14 | 24.97 | 6.76E+04 |
| 15 | **24.14** | 1.2E+05 | 15 | 24.38 | 1.01E+05 |
| 16 | 22.55 | 3.6E+05 | 16 | 22.88 | 2.84E+05 |
| 17 | 24.00 | 1.3E+05 | 17 | 24.81 | 7.54E+04 |
| 18 | 25.30 | 5.4E+04 | 18 | 28.33 | 6.71E+03 |
| 19 | 25.50 | 4.7E+04 | 19 | 25.45 | 2.39E+05 |
| 20 | 26.27 | 2.8E+04 | 20 | 29.46 | 2.27E+05 |
| 21 | 25.54 | 4.6E+04 | 21 | 25.95 | 2.27E+05 |
| 22 | 25.87 | 3.7E+04 | 22 | 27.42 | 4.79E+05 |
| 23 | **24.04** | 1.3E+05 | 23 | 24.45 | 5.42E+04 |
| 24 | 25.24 | 5.6E+04 | 24 | 25.20 | 6.76E+04 |
| 25 | 29.70 | 2.6E+03 | 25 | 25.07 | 1.01E+05 |
| 26 | 25.47 | 4.8E+04 | 26 | 26.32 | 2.84E+05 |
| 27 | 25.14 | 6.0E+04 | 27 | 26.12 | 7.54E+04 |
| 28 | 27.14 | 1.5E+04 | 28 | 27.41 | 6.71E+03 |
| 29 | 27.15 | 1.5E+04 | 29 | 27.34 | 1.33E+04 |
| 30 | 28.86 | 4.7E+03 | 30 | 27.24 | 1.42E+04 |
| 31 | 25.27 | 5.5E+04 | 31 | 26.24 | 1.42E+04 |
| 32 | 26.44 | 2.5E+04 | 32 | 26.64 | 2.14E+04 |
| 33 | 28.96 | 4.4E+03 | 33 | 28.92 | 4.47E+03 |
| 34 | 27.89 | 9.1E+03 | 34 | 27.82 | 9.53E+03 |
| 35 | 27.04 | 1.6E+04 | 35 | 26.66 | 2.12E+04 |
| 36 | 28.13 | 7.7E+03 | 36 | 27.05 | 1.62E+04 |
| 37 | 30.54 | 1.5E+03 | 37 | 30.13 | 1.95E+03 |
| 38 | 28.14 | 7.6E+03 | 38 | 29.36 | 3.31E+03 |
| 39 | 29.76 | 2.5E+03 | 39 | 30.12 | 1.96E+03 |
| 40 | 27.65 | 1.1E+04 | 40 | 28.19 | 7.39E+03 |
| 41 | 30.13 | 1.9E+03 | 41 | 30.14 | 7.39E+03 |
| 42 | 28.43 | 6.2E+03 | 42 | 29.48 | 3.04E+03 |
| 43 | 31.05 | 1.0E+03 | 43 | 31.61 | 7.04E+02 |
| 44 | 29.24 | 3.6E+03 | 44 | 29.51 | 2.98E+03 |
| 45 | 30.10 | 2.0E+03 | 45 | 31.19 | 9.40E+02 |
| 46 | 31.54 | 7.4E+02 | 46 | 31.34 | 8.48E+02 |
| 47 | 35.19 | 6.0E+01 | 47 | 34.55 | 9.34E+01 |
| 48 | 32.06 | 5.2E+02 | 48 | 31.19 | 9.40E+02 |
| 49 | 35.22 | 5.9E+01 | 49 | 36.04 | 3.35E+01 |
| 50 | 36.36 | 2.7E+01 | 50 | 35.83 | 3.87E+01 |

CT values given in bold indicate that SARS-CoV-2 could be propagated in cell culture.

**Table 2:** RDTs evaluated in this study

| # | Name | Manufacturer/distributor | # of evaluating laboratories |
| --- | --- | --- | --- |
| Classical Point-of-Care tests |  |  |  |
| 1 | SARS-CoV-2 Rapid Antigen Test | SD BIOSENSOR, distr. by Roche Diagnostics GmbH | 6 |
| 2 | Panbio™ COVID-19 Ag Rapid Test Device (Nasopharyngeal) | Abbott | 5 |
| 3 | RIDA® QUICK SARS-CoV-2 Antigen | R-Biopharm AG | 3 |
| 4 | Cleartest | servoprax GmbH | 2 |
| 5 | Dedicio COVID-19 Ag plus Test | Jiangsu Changfeng Medical Industry Co., Ltd., distr. by nal von minden GmbH | 3 |
| 6 | NADAL® COVID-19 Ag Test | nal von minden GmbH | 3 |
| 7 | BIOCREDIT COVID-19 Ag | RapiGEN Inc., distr. by Weko Pharma | 3 |
| 8 | STANDARD™ Q COVID-19 Ag Test | SD BIOSENSOR | 2 |
| 9 | BIOSYNEX COVID-19 Ag BSS | Biosynex Swiss SA | 2 |
| 10 | MEDsan® SARS-CoV-2 Antigen Rapid Test | MEDsan GmbH | 2 |
| 11 | NowCheck® COVID-19 Ag Test | BIONOTE, distr. by concile | 3 |
| 12 | COVID-19 Rapid Ag Test Cassette | Zhejiang Orient Gene Biotech Co., Ltd | 2 |
| 13 | ESPLINE® SARS-CoV-2 | Fujirebio Inc. | 2 |
| 14 | COVID-19 Ag Respi-Strip | Coris BioConcept | 2 |
| 15 | Healgen Coronavirus Ag Rapid Test Cassette (Swab) | Healgen Scientific LLC | 2 |
| 16 | Keul-o-test® | Günter Keul GmbH | 2 |
| 17 | Acro Rapid Test COVID-19 Antigen Rapid Test (Nasopharyngeal Swab) | Acro Biotech. Inc. | 2 |
| 18 | COVID-19 Antigen Schnelltest (Nasen-Rachenabstrich) | MEXACARE GmbH | 2 |
| 19 | COVID-19 Antigen Rapid Test Kit | Beijing Beier Bioengineering Co., Ltd. | 2 |
| 20 | mö-screen Testkit Corona Antigen Nasenabstrich | möLab GmbH | 2 |
| 21 | Medicovid SARS-CoV-2 AG Antigen Schnelltest | Xiamen Biotime Biotechnology Co., Ltd., distr. by MEDICE Arzneimittel Pütter GmbH & Co. KG | 2 |
| 22 | GENEDIA COVID-19 Ag | Green Cross Medical Science Corp. | 2 |
| 23 | COVID-19 Antigen Test Kit | Guangdong Wesail Biotech Co., Ltd., distr. by Bio-Gram Diagnostics GmbH | 2 |
| 24 | COVID-19 Antigen Schnelltest (Colloidal Gold) | Joinstar Biomedical Technology Co., Ltd., distr. by care impuls Vertriebs GmbH | 2 |
| 25 | blink COVID-19 Antigen Rapid Test (Nasopharyngeal Swab) | Hangzhou Realy Tech Co. Ltd., distr. by TREKSTOR GmbH | 2 |
| 26 | Novel Coronavirus (COVID-19) | Hangzhou Laihe Biotech Co., Ltd., | 2 |
|  | Antigen Test Kit | distr. by Lissner Qi GmbH |  |
| 27 | COVID-19 Antigen Rapid Test Strip | Koch Biotechnology (Beijing) Co., Ltd. | 2 |
|  | RDTs requiring a read-out device |  |  |
| 28* | STANDARD™ F COVID-19 Ag FIA | SD Biosensor | 4 |
| 29* | Sofia SARS Antigen FIA | Quidel Corporation | 3 |
| 30* | ScheBo® SARS-CoV-2 Quick™ Antigen | Schebo Biotech AG | 3 |
| 31* | Wantai SARS-CoV-2 Antigen Rapid Test (FIA) | Beijing Wantai Biological Pharmacy | 2 |

RDTs marked with \* require a test-specific device and are not to be judged visually with the device.

### RDT selection

RDTs included in this study were selected according to availability at the time of the study (table 2). No technical assumptions were made in the RDT selection process.

### RDT procedure

In general, RDTs were used as recommended by the manufacturer’s instructions. 50 µL of each pool were either subjected to the RDT using all buffers in volumes as recommended by the manufacturer, or the swab included in the RDT kit was used to absorb the 50 µL of each pool and then subjected to the RDT procedure as recommended by the manufacturer. Results obtained by visual examination of the test device by different laboratories were categorized as “positive” or “negative” and subjected to statistical analysis by using the GraphPad Prism software as indicated in the results section. Results were only accepted when the control band was positive, which was the case in more than 99% of the tests run.

## Results

### Evaluation panel establishment

Prior to distribution of the panel we compared the detectability of specimens before and after freezing at −40°C with two RDTs (#2, #3) and observed no significant differences in the virus concentration range close to the detection limit (data not shown). Furthermore, to test whether pooling and freezing had an impact on the detectability of specimens, we compared up to 44 fresh clinical specimens, representing a CT value range from 20 to 35 (1.8×10^7^ to 7.0×10^3^ genome copies per mL), with the 32 pools of Panel 1V1 covering the same range with in total 10 RDTs (#2, #3, #7, #8, #10, #11, #14, #21, #28, #31). In none of the RDTs tested we observed a significant discrepancy between the detectability of fresh specimens and pools with a similar CT value (data summarized in supplemental figure 1).

Supplemental figure 1: Comparison of 10 randomly selected RDTs with pools from Panel 1V1 (n≤32) and fresh clinical specimens (n≤44), covering a genome load from approximately 10^7^ to 7×10^3^ genome copies per mL. None of the RDTs showed a significant difference in the detectability of pools in comparison to fresh specimens.

### RDT analytical sensitivity

The panel was shipped on dry ice to the participating laboratories. Prior to use, the panel was thawed, mixed and aliquoted, and identical aliquots were used immediately to allow maximum comparability between laboratories and test days. Figure 1 summarizes the workflow as recommended to the participating laboratories. Assessment of results was performed visually by experienced laboratory personnel.

**Figure 1:**
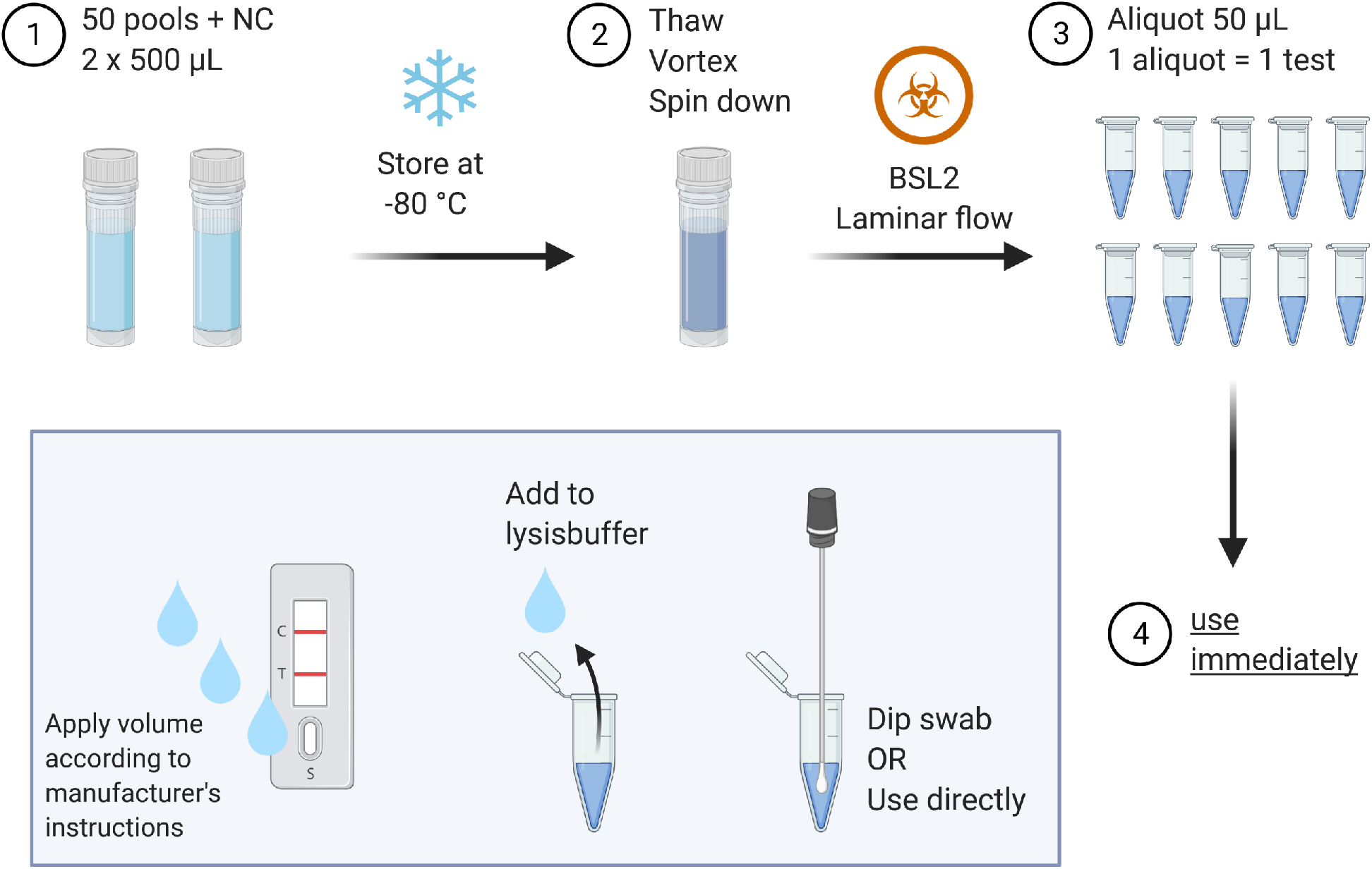
Recommendation for panel usage to guarantee maximum comparability between different laboratories and points in time of validation.

Figure 2 comprises the results obtained for the sensitivity of 27 RDTs that could be analysed visually (RDT #1–27) and the results for RDTs that needed a device for read-out (RDT #28– 31). Based on binary logistic regression of merged results obtained from different laboratories, the 50% probability to detect a certain genome load was calculated as a marker for the detection limit of an RDT. Six out of 31 RDTs showed a 50% detection probability for genome loads higher than 10^6^ genome copies per mL. Out of 31 RDTs, 24 had a 50% detection probability of less than 10^6^ genome copies per mL, while 15 had a 50% detection probability of less than 10^5^ genome copies per mL. The most sensitive RDTs detected approximately 75,000 genome copies per mL with a probability of 90%, while the least sensitive RDT showed a 90% detection probability for 2.3 × 10^7^ genomes per mL.

**Figure 2:**
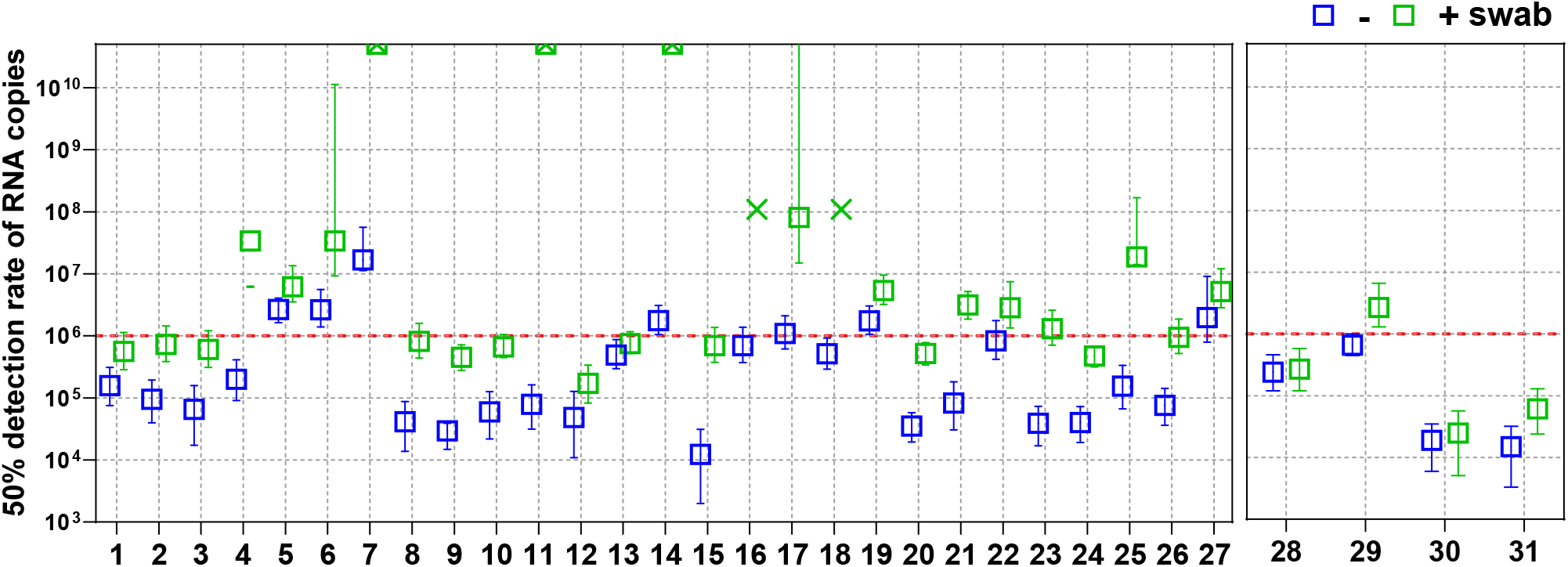
Analytical sensitivity expressed by the 50% detection probability of 31 RDTs. RDTs 28 to 31 require an additional device for reading out the result. Binary logistic regression was applied to calculate the analytical sensitivity from the 50 samples included in the panel. Open blue bars represent the results when virus-containing pools were directly subjected to the RDT without using a swab. Open green squares represent the corresponding results when using a swab. Crossed squares indicate that no results were determined with swabs, while green crosses show the one positive result from a pool with the corresponding virus load to CT 17.55, pool #1.

Figure 2 also shows that using the RDT-specific swabs to absorb the pool material prior to testing can lead to a loss of analytical sensitivity of approximately the factor 10 to 50, although some of the RDTs showed only small differences between direct application of the pool and swab usage. This indicates a varying efficiency of the absorption and release characteristics for SARS-CoV-2 particles from the swabs.

### RDT clinical sensitivity

To determine the clinical sensitivity of an RDT, results of different laboratories were merged and categorized according to the genome load, e.g. CT value obtained for each pool by real-time PCR [8a]: CT<25 (10^6^ genome copies per mL), 25<CT<30 and CT>30. Sensitivities of each RDT to identify pools correctly were calculated for each CT category.

According to previous studies, CT<25 corresponds to an increased probability of a specimen to successfully propagate SARS-CoV-2 in cell culture [9–11]. This was confirmed for 9/18 pools with CT values <25 for Panel 1V1 and 5/17 pools for Panel 1V2. Pools with a CT>30 were highly unlikely to contain virus amounts high enough to grow in cell culture. Specimens with CT values between 25 and 30 very rarely propagated virus in cell culture.

Figure 3 summarizes the sensitivities for the 31 RDTs evaluated. Blue circles represent results when no swab was used and green circles those when swabs were used. Figure 3A shows the sensitivity for the whole number of 50 SARS-CoV-2-positive pools of the panel. Results are further categorized according to the different virus loads represented by a pool. Figure 3B covers pools with CT<25 (n=18 Panel 1V1, n=17 Panel 1V2, potentially infectious). Figure 3C shows pools with CT>25 (n=32 Panel 1V1, n=33 Panel 1V2) and figure 3D pools with a CT value between 25 and 30, which is the range where the RDTs showed significant differences (n=23 for both panels). The number of laboratories contributing to a result is given in table 1.

**Figure 3:**
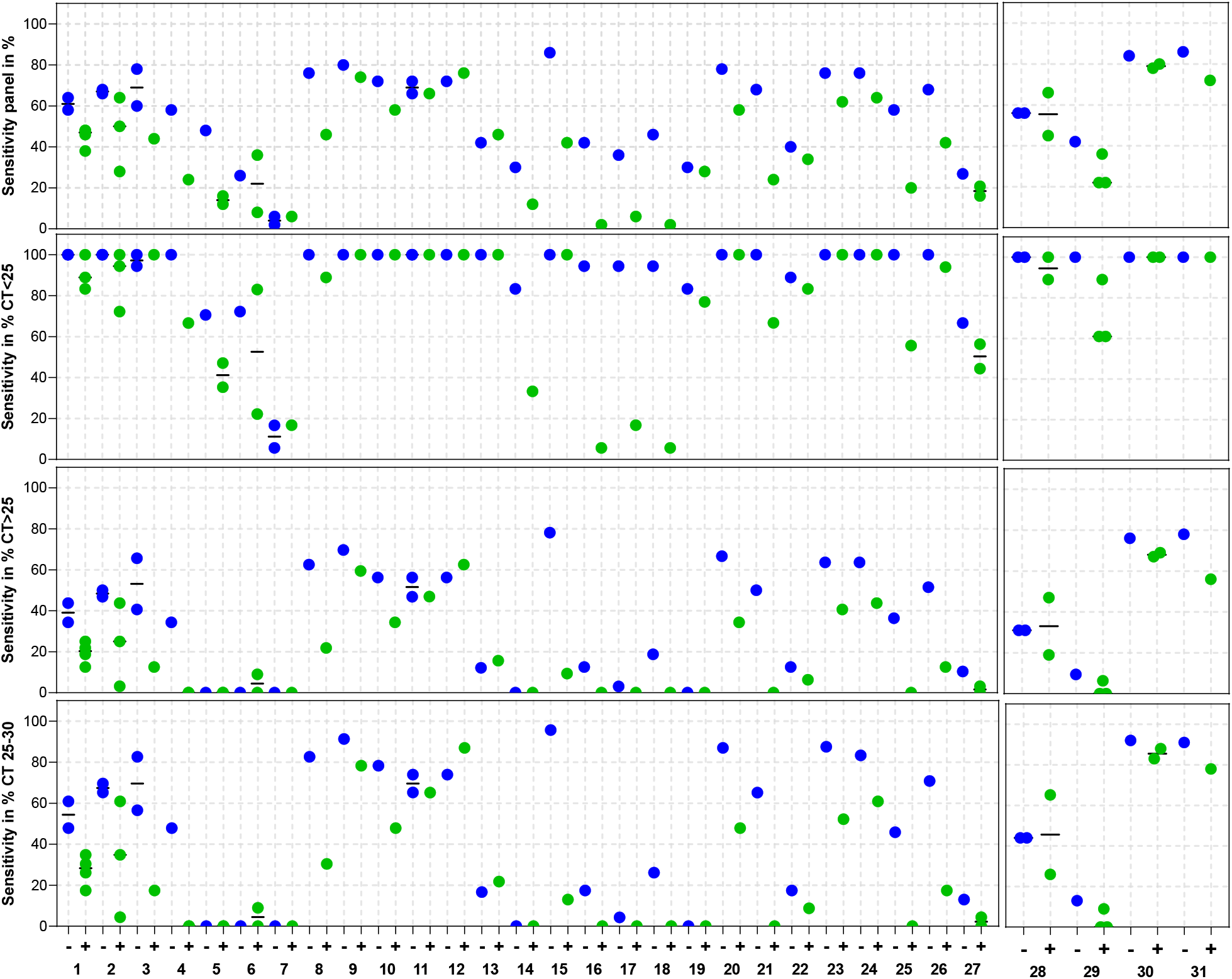
Clinical sensitivities of 31 RDTS as determined by two to six laboratories using 50 pools from evaluation Panels 1V1 and 1V2. Blue circles indicate direct application of the pool to the test buffer and green circles indicate results using the RDT-specific swab. Each circle represents the results of one laboratory (symbols may cover each other). RDTs 28 to 31 require an additional device for reading out the result.

The data reveal that for virus loads higher than 10^6^ genome copies per mL (CT<25) the sensitivity of 26/31 RDTs was higher than 80%, indicating that these RDTs would potentially identify infectious specimens with a probability of 80% (figure 3A). For the latter RDTs, the proficiency to detect these pools that contain culturable SARS-CoV-2 was even better with values up to 100% (data not shown). For a virus load <10^6^ genome copies per mL (CT>25), none of the evaluated tests could surpass a sensitivity of 80% (figure 3B). In the range between CT 25 to CT 30, 10/31 RDTs reached a sensitivity of 80% and higher, even if five further RDTs showed sensitivities close to 80% (figure 3D). Finally, when the sensitivity to detect all 50 pools of the panel was determined, only 4/31 RDTs passed a sensitivity of 80% or higher, with two of the four RDTs requiring a detection device for read-out. However, 10/31 RDTs showed an overall sensitivity higher than 70% (figure 3A). Using a swab to reduce the volume of the pool material in the RDT test procedure can lead to reduced sensitivity due to loss of virus particles in the absorption and release process. Hence, as described for the analytical sensitivity based on the RNA detection limit, clinical sensitivities were lower for most RDTs when a swab was used.

Even if most of the RDTs were analysed in two independent laboratories only, RDTs 1–5 and 28–30 were evaluated 3–6 times with or without using swabs. As shown in figures 2 and 3, there can be significant variability for some tests, which is most likely due to the more subjective interpretation of a positive signal; however, most results were very similar even when generated in different laboratories on different days.

## Discussion

RDTs are promising tools in the diagnostic portfolio of tools for the identification of SARS-CoV-2-infected individuals [13–15]. Since these tests do not use amplification of the target, like PCR, their analytical sensitivity is usually limited. Hence, the evaluation of RDTs plays a major role in defining the proper applications of RDTs. In contrast to PCR, where the specimen can be inactivated, RDTs should be evaluated with clinical material that contains native viruses to mirror the diagnostic application as authentically as possible. However, the systematic comparison of various RDTs in different laboratories at different times makes larger sample volumes and a good storage stability necessary.

Multiple sampling of naso- and/or oropharyngeal swabs is hampered by reproducibility. Even sampling the same patient with several swabs consecutively will likely lead to a variation in the virus load per swab which has to be controlled by real-time PCR, changing the test procedure. Because most of the RDT protocols require the clinical specimen to be sampled with a swab from which virus has to be eluted in the system-specific buffer, one swab cannot be used more than once without changing the protocol. Therefore, the decentralized evaluation of various RDTs in different laboratories is obviously difficult. So far, clinical samples with semi-quantified SARS-CoV-2 concentrations or virus propagated in cell culture have been used to validate RDTs [16–18]. This can be done for a limited number of tests in a short period of time but is not suitable when numerous RDTs have to be compared regularly in different laboratories.

Therefore, we established an evaluation panel that was used to determine the analytical and clinical sensitivity of RDTs providing comparable results. The main basis of each pool were dry swabs in PBS originally used for PCR diagnostics. Since some of the swabs were transported to the laboratory in medium, the final concentration of medium in pools was ≤20% for each pool, ≤10% for 38 pools and 0% for 25 pools. Ten RDTs were randomly selected to calculate whether the medium content had an impact on the sensitivity. As shown in supplemental figure 2 for one of the RDTs, binary logistic regression revealed no difference in the detection probability of pools containing varying amounts of medium. Although these results are only derived from ten RDTs, we believe that this low medium content has no influence on the test sensitivity in general, since pools with medium percentages between 10% and 20% were distributed over the whole panel 1V1.

Supplemental figure 2: Impact of medium contents in the 50 pools included in Panel 1V1. Binary logistic regression was used to demonstrate that the amount of medium ranging from 0% to 20% has no impact on the detection limit of one randomly selected RDT.

SARS-CoV-2 genome load was determined by real-time PCR in clinical specimens, and those specimens with similar load were pooled diluted in a background of negative swabs and the virus load quantified again. The established pools had a volume of 10 mL for both panel 1V1 and 1V2 and covered a genome load from 1.1×10^9^ genomes per mL down to 400 genomes per mL, which is the range of typical clinical specimens analysed in our laboratory. Even if the genome load does not reflect the number of virus particles directly, the RNA copy number was recently used to estimate the number of virus particles reflecting the infectious potential of a specimen and can correlate with the N protein concentration in clinical samples [19].

The fact that we used pools of up to 10 clinical specimens facilitates to compensate to some degree for a potential variation between individual samples, for example varying ratios of genome copies to the number of viral particles or rather the antigen concentration.

We could demonstrate that the pools of the panel showed results comparable to those for fresh clinical specimens with similar SARS-CoV-2 genome load when selected RDTs were used, and that freezing at −40 to −80°C did not impact the detectability significantly. Nevertheless, as a trade-off for better reproducibility and comparability, the material used was not as fresh as in a clinical setting [.

Usually, RDTs use swabs that are subjected to the RDT-specific buffer before incubation of the test membrane is done. Our approach necessarily began with liquid specimens of 50 µL, which is intended for some RDTs, but not for all of them. However, with the intention to allow the generation of comparable and reproducible data, we accepted that some of the buffers were diluted by the fluid of the pools. Adding 50 µl of pools directly to three randomly selected RDTs without additional test-specific buffer led to results indicating that additional buffers were not required at least in these RDTs.

The strategy of using liquid specimens comes with a further benefit, e.g. the option to cultivate the pools in cell culture, showing the infectivity of pools with a sufficient virus load. To our knowledge, this is one of the few studies that systematically evaluated several commercially available Ag RDTs using standardized samples in comparison to qPCR as well as infectivity data from cell culture [21]. However, since all specimens included in a pool have been frozen and thawed at least once, the capability to grow in cell culture can be improved with fresh clinical specimens of a comparable virus amount [22].

Variability observed in different laboratories has been described for diseases like malaria [23] and was significant for some RDTs, but in general highly comparable results were obtained. Therefore, our results reflect the natural variance that can be expected when different users apply the RDTs. Over time we observed that users adopted the test and that the differentiation of weak positive results from negative results became better with increasing experience. Therefore, it can be assumed that the interpretation of results is better standardized with RDTs that require a device to read out the signal. Based on the number of RDTs we have validated, we can confirm that read-out devices can help generate better reproducible results and reduce the inter-user variance.

### Influence of swabs on the sensitivity of RDTs

The common RDT test starts with the sampling that results in a swab containing material from the mucosa of the naso- or oropharynx. Then the swab is put into the RDT-specific buffer and is subjected to the test membrane. Applying liquid evaluation specimens with known virus amounts therefore does not consider the impact of the swab on the result. The swab has to absorb liquids from the mucosa potentially containing SARS-CoV-2 or can scrape cellular material containing virus; probably it is a combination of both. Beside the problem that the swab does not quantitatively absorb the specimen, virus proteins can stick to the swab and will not be subjected to the RDT, reducing the analytical sensitivity. While some of the RDTs do not suffer significantly from using a swab prior to testing (#13, #28, #30), most of the tests lose sensitivity approximately by the factor 10 to 20. This does not mean that the respective test device is inferior to the previously mentioned tests, but rather that the swab is not efficient in absorbing and releasing SARS-CoV-2 from a liquid specimen. However, speculating that these swabs will come with the same drawbacks when used in clinical sampling, the loss of sensitivity can also occur. In a patient carrying for example 10^6^ genome copies in the nasopharynx, with RNA load used as surrogate for viral particles, the sampling on the mucosa bears the risk to obtain a false negative result in the RDT due to considerable loss of antigen in the swab. Further investigations of the efficiency of virus absorption and release from a swab will help interpret the risk of false negatives by sampling. Besides, further specimens like saliva are under investigation for their use in RDTs.

Finally, our study did not address the specificity of an RDT and cannot assess the risk of false positive results. However, recent studies show that high specificity is reached by most of the RDTs evaluated [24]. In addition to SARS-CoV-2-positive specimens, the next version of the evaluation panel will also include specimens containing SARS-CoV-2-negative specimens positive for further respiratory viruses.

## Conclusion

The sensitivity of the 31 RDTs evaluated in this study varied significantly and depended largely on the virus load in the respective specimen. While four RDTs showed a sensitivity of >80% over the whole range of virus loads investigated, 26 RDTs had a sensitivity of >80% for potentially infectious specimens, indicating that sensitive RDTs can be used to identify contagious individuals in various settings, but not to identify infected individuals with lower virus loads. Our results are in agreement with several other studies not using a standardized evaluation panel [25], indicating the applicability of the described panel for RDT evaluation. The minimal performance characteristics for an RDT have been recently discussed to be at least 80% for symptomatic patients [13]. Considering varying virus loads during the time course of infection in an individual and in between individuals, the sensitivity of an RDT should therefore better be attributed to a certain virus load instead of the time after onset of symptoms or a qualitative PCR result.

## Data Availability

All data are published and hence available.

## Acknowledgements

The authors are grateful to Ursula Erikli for copy-editing. This work was funded by the Ministry of Health, Germany (Maßnahmepaket 1&2).

## Authors contributions

Andreas Puyskens, Eva Krause, Janine Michel and Marica Grossegesse designed the study, established the evaluation panel and performed the RDTs with Roman Valusenko. Daniel Bourquain performed cell culture experiments. Micha Nübling and Heinrich Scheiblauer characterized the panel and evaluated several RDTs. Viktor Corman, Christian Drosten, Katrin Zwirglmaier, Roman Wölfel, Constanze Lange, Jan Kramer, Johannes Friesen, Ralf Ignatius, Michael Müller, Jonas Schmidt-Chanasit, and Petra Emmerich provided data for varying numbers of RDTs with the evaluation panel. Lars Schaade and Andreas Nitsche conceptualized the study, analysed the data and wrote the manuscript.

