## Supplementary figures and images for "Establishment of an evaluation panel for the decentralized technical evaluation of the sensitivity of 31 rapid detection tests for SARS-CoV-2 diagnostics"

### Supplemental Figure 1

#### Supplemental figure 1

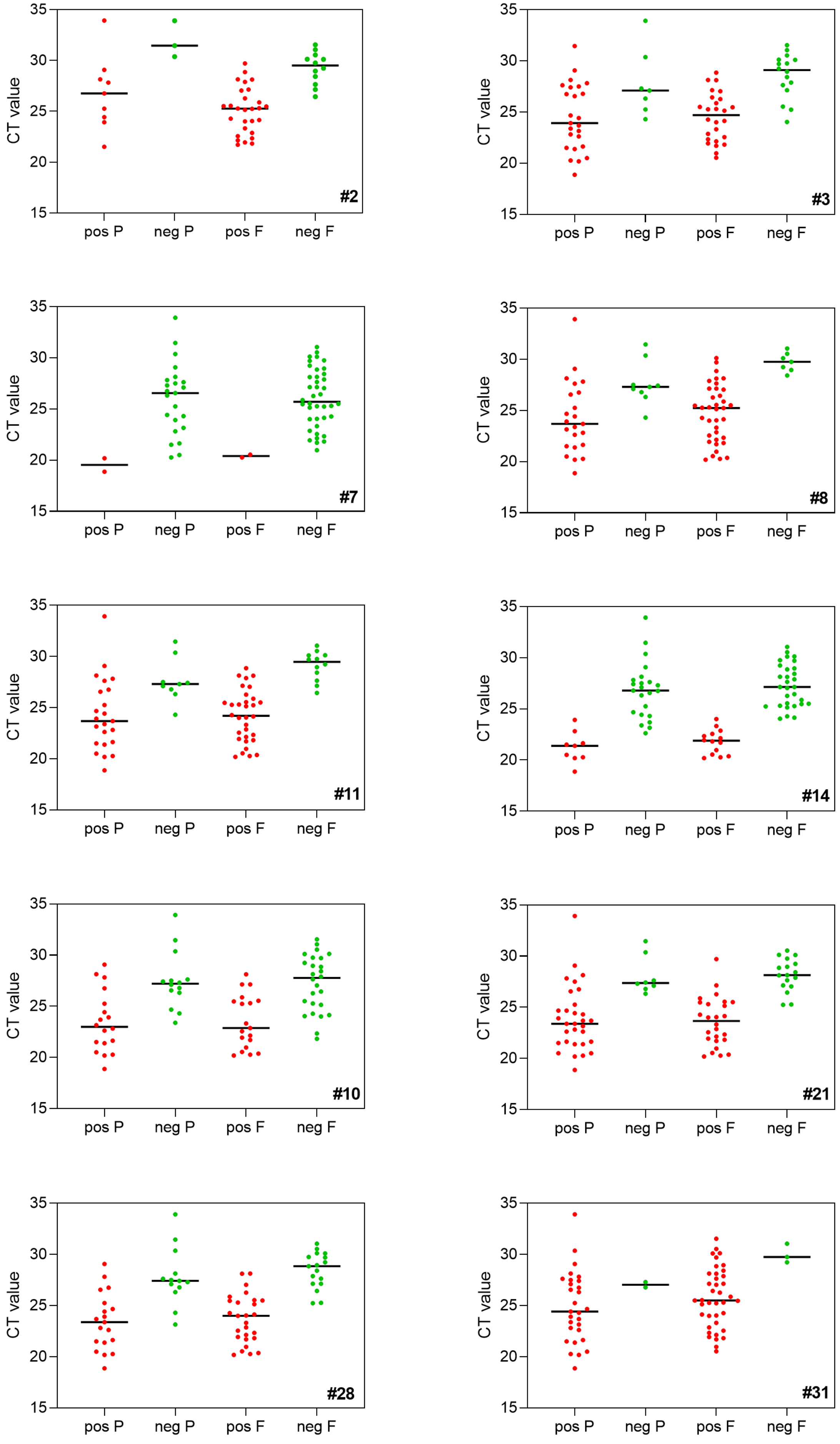

### Supplemental Figure 2

Supplemental figure 2

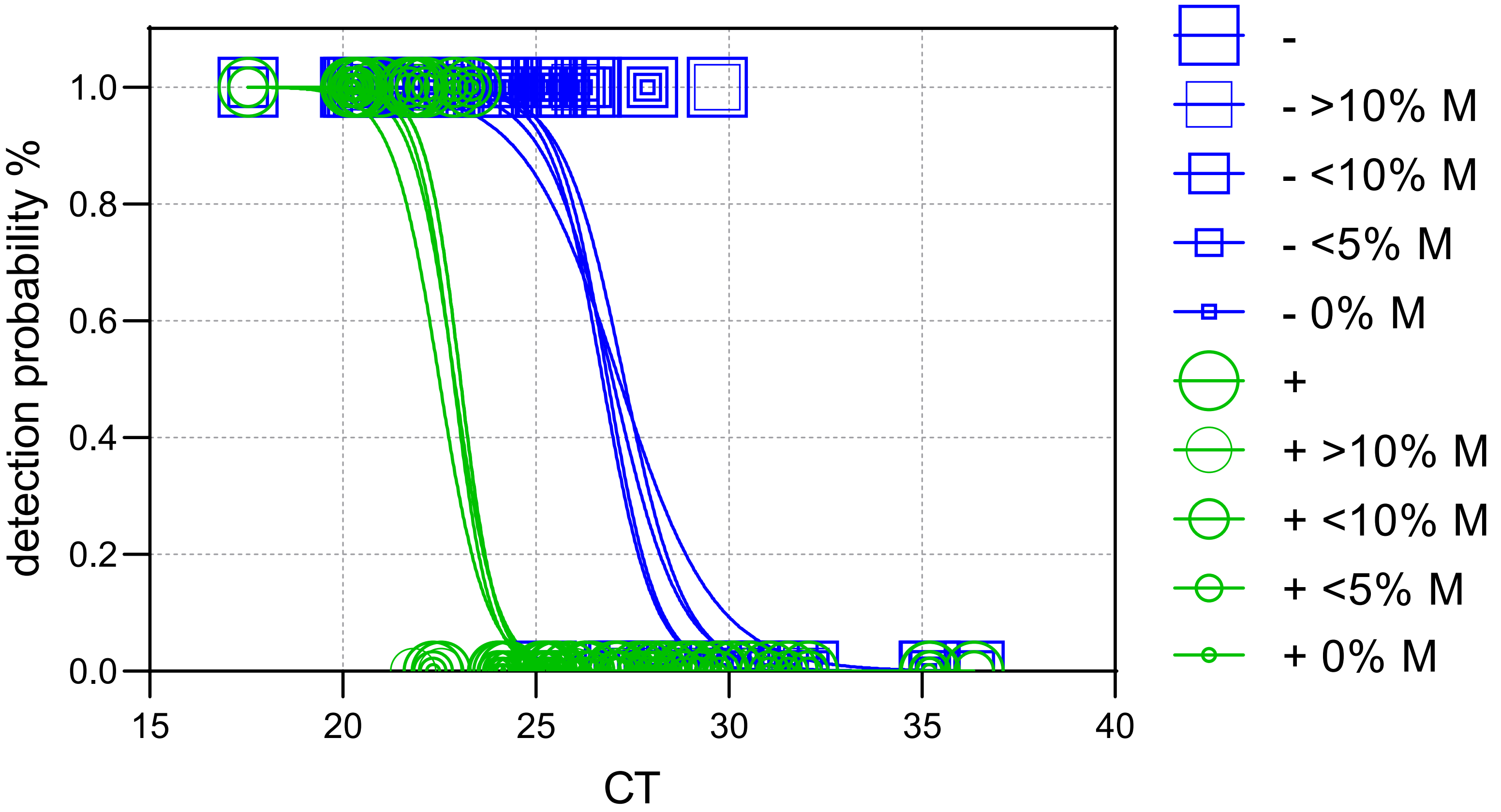
